# A quest for the origin of the uneven spread of Covid-19 cases

**DOI:** 10.1101/2021.07.14.21260550

**Authors:** Arunava Bhadra, Mahasweta Bhattacharya

## Abstract

For more than one and a half years now the world is highly impacted by the Covid-19 pandemic. The covid-19 cases are, however, not evenly distributed across the countries; a few countries, particularly high-income countries have been hit harder than the countries of weak economic condition. The reasons for such an asymmetrical distribution are not clearly understood yet. In the present work, we have examined the unevenness of global distribution of Covid-19 incidences till 18^th^ June 2021 in terms of the economic condition of countries. Subsequently, we have tried to identify the main underlying factors behind unequal Covid-19 cases. Our analysis suggests that the degree of Isolation, the diffusion of coronavirus (interconnectivity), and the percentage of elderly people in the population are the main causes of the different unequal spread of Covid-19 cases in different countries. We find that the Covid-19 infected and death cases are well describe by a power law in terms of the stated parameters.

## 1, Introduction

Since its emergence at the end of 2019, the Covid-19 pandemic has affected billions of people worldwide including the loss of life of around four million till now. Great efforts have been made so far to understand the disease, which is caused by Severe Acute Respiratory Syndrome Coronavirus 2 (SARSCOV-2), and also to develop mechanisms for fighting with it as well as to prevent from the disease but on various aspects our knowledge about the disease is still limited.

The severity of the Covid-19 pandemic is not evenly distributed around the geographical locations (Johns Hopkins Coronavirus Resource Center 2021, Rodríguez-Pose 2021, The 2021). Even within a country or even within a city, some locations recorded significantly higher infection and mortality compared to other places (Huang 2020, Hohl, 2020, Sun 2021, Yang 2021). The timing of occurrence and severity are also not the same for all the countries/states. To understand the cause of such variation a large number of studies have been made to explore the dependence of Covid-19 infected and mortality rate on various environmental, demographic, health infrastructure, government response, and other parameters, see for instance (Bhadra 2020, Angel 2020, Sarmadi, 2020, Duhon 2021, Ma 2021). Though some order of positive correlation on a few parameters for a few countries is noticed [8] which are not sufficient to resolve the observed geographical inequality of Covid-19 incidences.

The Covid-19 pandemic so far has also exhibited some unexpected behavior. For instance, a large share of the reported infection and death toll is found in the high-income (developed) countries (Schellekens 2020, Mukherjee 2021, Chatterjee 2021). In contrast, the Africa continent has registered significantly less number of Covid-19 infection and mortality despite weaker financial and healthcare conditions. It was stated in (Schellekens 2020) whether the world has two Covid pandemics. However, several social and demographic parameters depend on economic conditions. For instance, the median age of a country is strongly correlated with the economical well-being of the residence of the country (Schellekens 2020). In a few previous analyses, it was demonstrated that the difference in the age distribution of lower-income and higher-income countries is an important cause of higher Covid-19 related mortality in high-income countries (Schellekens 2020, Mukherjee 2021). Also, under-reporting of Covid-19 infected cases due to poor testing capacities has been mentioned as another important cause (Schellekens 2020). Other probable causes as proposed by different researchers include air quality (Kapitsinis 2020, Coccia 2021) global interconnectedness (Kapitsinis 2020,) expenditure towards healthcare infrastructure (Coccia 2021), prevalence of autoimmunity (Chatterjee 2021) etc.

In recent times a substantial number of Covid-19 related infections and mortality have been noticed in the middle and lower-middle-income countries like Brazil, Mexico, Ecuador, India, Russia, and a few others. So it is imperative to re-inspect the prevailing perception of high Covid-19 disease in high-income countries and look for a better clue about uneven distribution. With this objective in the present work, we shall first examine that to what extent the spread of Covid-19 cases is uneven among the countries/region of different economic conditions at present. Subsequently, we shall explore the origin of the unevenness in Covid-19 spread depending on the economic condition. Because of the contagious nature of Covid-19, we particularly focus on a couple of variables that reflect the degree of isolation and interconnectivity. Besides, the association of Covid-19 cases with the elderly population will be examined.

The organization of the paper is as follows. In the next section, we shall discuss the data used, the source of the data, etc. In section 3 we briefly mention the analysis technique adopted in the present work. In section 4 we shall present the results of the present analysis. We discuss our results in section 5 and conclude in the same section.

2, Data used: In this work, we have used worldwide country-wise cumulative Covid-19 infected and mortality cases as on 18.06.2021. The Covid-19 data is taken from the world health organization website (https://covid19.who.int/table) where the Covid-19 data for 196 countries including Holy see, and 42 islands, territories, and others are figured. To avoid the probable large statistical fluctuations in the data we have not included the countries with population less than 0.5 million (the cut-off is set in an arbitrary manner) and all the 42 territories/islands/other.

The Gross National Income (GNI) is often used as standard measures of the financial state of a nation. Here we have used GNI per capita in Purchasing Power Parity to neutralize the effect of the net population of a country. The GDP and GNI data for the countries are taken from World Bank repository data (https://data.worldbank.org/). The data for a few countries like North Korea are not available and hence we could include those countries in our analysis.

We have also used the demographic data of the countries under study. We have particularly considered the country-wise data of population, life expectancy at birth, median age and percentage of the urban population, and total air passengers in a calendar year. We have taken the data from the World Bank (https://data.worldbank.org/) and the worldometer (https://www.worldometers.info/demographics/). The Covid-19 testing data has been taken from the worldometer.

We get the required data for a total of 165 countries except for the total air passenger data which have been included in the present analysis. The total air passenger data of only 141 countries are found out of the 165 countries for which all the other data are available.

### Analysis Methods

Our objective is to look for any correlation between the Covid-19 infected and mortality cases in a country with the economic condition and a few demographic and other parameters of the country which could be the probable underlying cause of unequal Covid-19 cases. To neutralize the population effect, we consider Covid-19 infected and death cases per million people. The GNI per capita in PPP has been used as the indicator of the economic condition of the citizens of a country. Regarding the demographic parameters, we have employed the median age, the percentage of population above 65 years in a country and the percentage of the urban population. To describe the interconnectivity we use the parameter the total air passenger in a year.

For the correlation analysis, we take the Covid-19 infected and mortality cases per million as the dependent variable. The weighted correlation coefficient is the standard measure for quantifying the statistical relationship between two variables, which is defined by

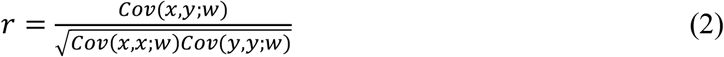

where 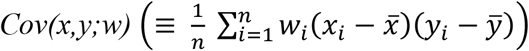 denotes the weighted cross-covariance of the two variables x and y and 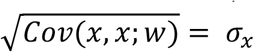 is the weighted standard deviation. Often each data points are treated equally so that weight factor wi is taken unity for all i. However, when the relative significance of each data points are not equal, one has to use the weighted correlation coefficient.

When the total variables are three with z as the dependent variable, we compute the multiple correlation coefficient which is defined as

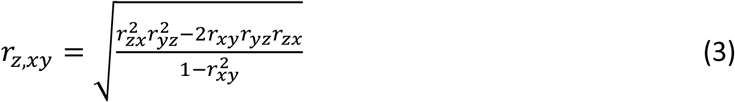

The strength of association between the dependent variable (z) and an independent variables (x)often evaluates by regression analysis. The equation of linear regression reads

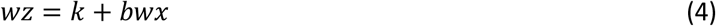

where k and the coefficient a are constants.

## 3. Results

Before making any analysis regarding an association between Covid-19 cases and any demographic and/or economic parameter it is prudent to ensure the data quality. Particularly, the issue of under-reporting has been ascribed by several researchers as one of the major reasons for low infected/death cases in some countries as already mentioned in the introductory section. To critically analyze this point we focus on the positivity rate as outlined below.

### 3.1 Positivity rate

The testing for Covid-19 is crucial for detecting people who are infected. The testing policy, however, differs among countries. It has been noticed that the countries having good healthcare systems and economically sound are testing a large section of the population, with or without any reported symptoms whereas many countries test only people having reported symptoms and/or those coming in contact with a covid-19 infected person. The question is that whether low testing effectively implies a large number of undetected Covid-19 infected people. This is a subtle issue. If there are no symptoms among the residents of a country, why shall they go for testing? The positivity rate (the ratio of the total number of detection for infection to a total number of testing) seems important in this regard. Though the positivity rate is a dynamic quantity but when a country is testing only people with a high prior probability of infection the positivity rate should be higher. In table 1 we present the positivity rate for different regions. The finding shows that though the testing is low in Africa the positivity rate is not high. The positivity rate of a few countries with a high total number of infected and mortality cases is shown in Table 2 for comparison. Here the data used still 7^th^ June 2021. Note that the testing data does not truly have one to one correspondence with the number of persons tested; often testing is done a number of times for a single person depending upon his/her health condition or because of other purposes such as (frequent) travel, joining a job/course, etc. So the results shown in Tables 1 and 2 should be considered as a good approximation only. It appears that in American countries, bearing USA, Canada, and a few others, there is a possibility of some under-reporting of the infected cases.

**Table 1:** Testing and positivity rate in different regions of the world

| Region | Testing per million |  | Positivity rate<br>(infected/testing) |  |  | Mortality/testing | Mortality rate |
| --- | --- | --- | --- | --- | --- | --- | --- |
|  | Mean | median | mean | median | Weighed mean |  |  |
| Africa | 74166 | 31556 | 0.103<br>±<br>0.018 | 0.072 | 0.104 | 0.0019 ± 0.0004 | 0.019 |
| America<br>(North and South) | 309044 | 218479 | 0.158<br>±<br>0.018 | 0.132 | 0.193 | 0.005 ± 0.001 | 0.035 |
| Eastern Mediterranean | 605412 | 176023 | 0.115<br>±<br>0.013 | 0.108 | 0.103 | 0.0044 ± 0.0012 | 0.032 |
| Europe | 1346615 | 922050 | 0.095<br>±<br>0.009 | 0.074 | 0.078 | 0.0021 ± 0.0003 | 0.023 |
| South-East Asia | 313778 | 111321 | 0.074<br>±<br>0.016 | 0.075 | 0.084 | 0.0010 ± 0.0003 | 0.015 |
| Western Pacific | 325787 | 131880 | 0.027<br>±<br>0.010 | 0.006 | 0.011 | 0.0003 ± 0.0001 | 0.037 |
| World | 579474 | 200974 | 0.1036<br>±<br>0.0070 | 0.079 | 0.085 | 0.0026 ± 0.0003 | 0.026 |

**Table 2:** Testing and positivity rate in a few countries

| Countries | Testing per million | Positivity rate |
| --- | --- | --- |
| USA | 1462915 | 0.068 |
| India | 263057 | 0.079 |
| Brazil | 232946 | 0.339 |
| France | 1329350 | 0.064 |
| UK | 2735997 | 0.024 |
| Mexico | 54776 | 0.341 |

The mortality rate, which is the ratio of total mortality to total infected cases of a country/region, may also be an important parameter to examine the under-reporting issue. In Table 1 we also give the mortality rate for the mentioned regions. The weighted average of mortality cases for the South Asian countries is found quite low whereas that for western Pacific and American countries are found comparatively higher. Since there is no effective medicine for Covid-19 yet the mortality rate depends mainly on the healthcare system of the country that includes the availability of Oxygen supply, hospital bed, number of medical staff (doctors, nursing and technical), etc. The high mortality for the western Pacific region is mainly due to China where the disease was detected first. At the early stage, when there was no idea how to treat the disease, the mortality rate was higher which is reflected in the mortality rate of China. The under-reporting of infected cases may be a reason for the higher mortality rate in South American countries. The Covid-19 pandemic arrived late in the south Asian countries compare to other regions except Africa and a prolonged lock-down imposed mainly in India may be the reasons for the lower mortality rate in the region. But even the mortality rate of African countries is around 0.02. There may be statistical fluctuations also in mortality rate but owing to quite large population in the south Asian countries and the limited healthcare facilities, particularly in the rural region, the possibility of under-reporting of death cases in the region by around 50% is not unlikely.

### 3.2 Association with the economic condition

The gross national income (GNI) includes the total income earned by all the residents of a country/region from home as well as from abroad and is generally considered as the main indicator of the net wealth of the country/region. The GNI per capita (per person) of a country/region arguably reflects the standard of living of the residents of the country/region. However, the price levels of commodities differ significantly from countries to countries which are accounted for by expressing GNI in purchasing power parity (PPP) to better describe the standard of living of the residents. Here we have considered GNI per capita in PPP as the economic parameter to assess any association of Covid-19 infection and death with the economic condition.

Figure 1 shows the variation of infected and death cases as a function of GNI/capita (PPP). There is a clear tendency that the infected cases increase with GNI/capita (PPP) which is not so pronounced for the Covid-19 related death cases. We divide the total population of the considered 165 countries in five groups (which is the maximum admissible since China and India individually contribute nearly 20% of the world population) according to their financial condition (GNI/capita in PPP) and estimate the Covid-cases in each group. The results are shown in Table 3. We note that the Covid-cases increase with average GNI/capita in PPP except for group 4 which is mainly contributed by China. Since the situation of China is an exception among the high populated countries, we re-grouped the countries excluding China and the Covid-cases in each group are shown in Table 4. It is found that here the Covid-cases consistently increase with the financial condition.

**Table 3:** Covid-19 cases, economic condition and related parameters in five groups from 165 countries based on the financial condition.

| Group | Fraction of the total population of the countries to Total population of all the countries | Average infected cases per million population | Average mortality cases per million population | Average GNI/capita in PPP in US\$ | Average percent of people live in Urban areas | Average % of people of age more than 65 years in the population | average Air passenger |
| --- | --- | --- | --- | --- | --- | --- | --- |
| 1 | 0.198 | 3131.3 | 59.8 | 3730 | 35.3 | 3.59 | 43.9 |
| 2 | 0.199 | 20690.6 | 267.6 | 6938 | 36.3 | 6.27 | 125.2 |
| 3 | 0.176 | 31354.8 | 932.1 | 12239 | 62.8 | 7.21 | 360.2 |
| 4 | 0.203 | 1367.4 | 18.1 | 16879 | 60.1 | 11.53 | 497.8 |
| 5 | 0.224 | 56832.7 | 1258.7 | 35865 | 80.0 | 16.23 | 1767.1 |

**Table 4:** Covid-19 cases, economic condition and related parameters in four groups of 164 countries (excluding China) based on the financial condition.

| Group according to financial condition | Fraction of the total population of the countries to Total population of all the countries | Average infected cases per million population | Average mortality cases per million population | Average GNI/capita in PPP in US\$ | Average percent of people living in Urban areas | Average % of people of age more than 65 years in the population | average Air passenger per year per 1000 population |
| --- | --- | --- | --- | --- | --- | --- | --- |
| 1 | 0.249 | 3094.8 | 59.1 | 3788 | 37.5 | 3.56 | 44.5 |
| 2 | 0.256 | 19810.4 | 255.9 | 7011 | 35.6 | 6.43 | 153.7 |
| 3 | 0.248 | 33691.8 | 1050.0 | 13949 | 66.6 | 7.71 | 602.2 |
| 4 | 0.246 | 58802.2 | 1191.3 | 47420 | 79.6 | 17.17 | 1749.4 |

**Figure 1.**
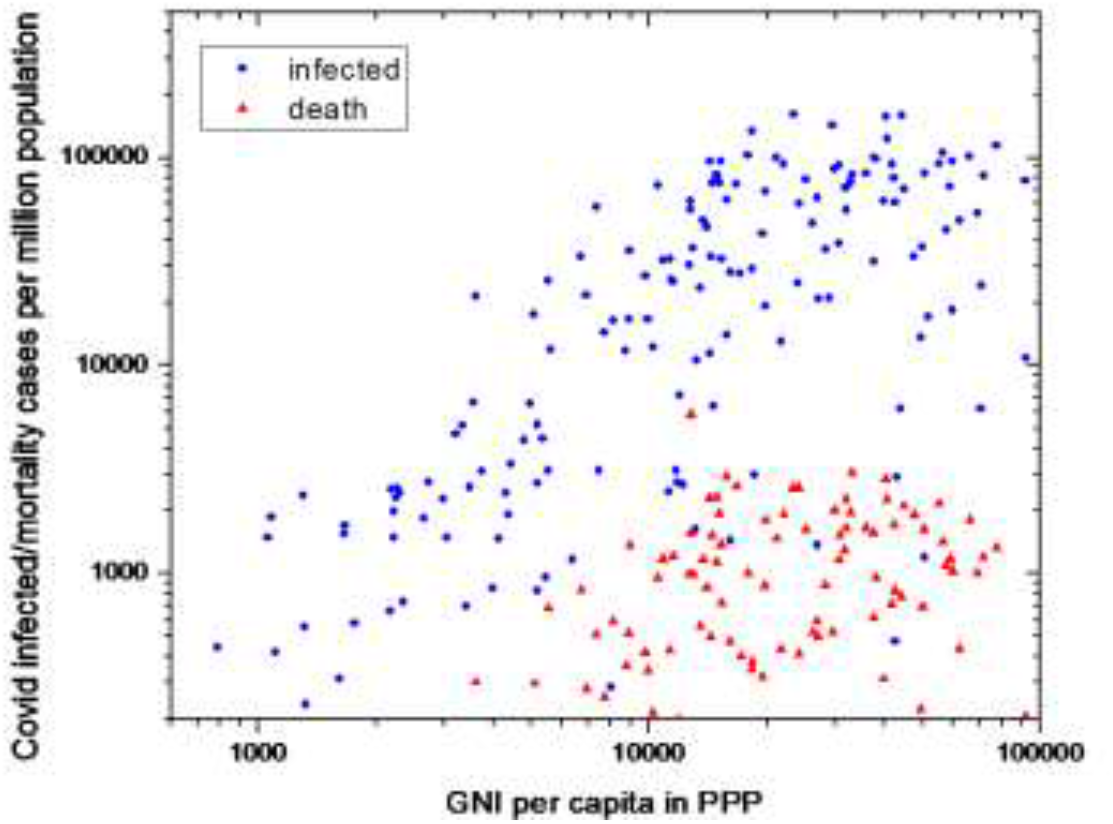
Variation of the Covid-19 infected/mortality cases per million population with the GNI/capita in PPP of total 165 countries world-wide.

We calculate the correlation coefficient (r) between Covid-19 infected and mortality cases per million people and the GNI per capita (PPP) of 165 countries without any weightage, and found r=0.49 (p = 7.7 × 10^−8^) and 0.30 (p = 2.9 × 10^−27^) for infected and mortality cases respectively, which shows a moderate to strong correlation between the rate of infected cases with the GNI per capita (PPP) but in the case of mortality, the correlation with GNI/capita (PPP) is weak to moderate. If GNI per capita is used without invoking PPP the r values become 0.38 and 0.20 respectively for infected and death incidences. The quoted p-values are from the t-test of the concerned data. In subsequent results, we shall not mention the p values because in all the cases we get very small values of p. In the above analysis, all the data points are weighted equally. Since the population of different countries differ significantly, the overall infected and mortality cases are not reflected correctly when equal weight for all the data points is adopted. Considering the fractional contribution of a country to the world population as the weight factor we have calculated the weighted correlation coefficient between Covid-19 cases and the GNI/capita in PPP and is found a stronger correlation r = 0.63 and 0.49 respectively for the infected and the death cases. The strong healthcare system of high-income countries seems responsible for lower correlation of death cases with financial conditions compare to that of the infected cases.

### 3.3 Covid-19 cases and the age structure of countries

The age distribution of the developed and the developing countries/region differs significantly. Life expectancy at birth varies widely among the countries, from 53.3 years in the Central African Republic to 84.4 in Japan. The average life span of the population is higher in economically rich countries than those in weaker economic conditions. Since the life span of residents of rich countries is longer, the population of elderly people in those countries is significantly higher than those in developing countries. As a result, the median age is higher in rich countries. Since elderly people are susceptible to any respiratory diseases, it may be a major cause of high Covid-19 cases in rich countries. To examine any association of Covid-19 cases with the age structure we have considered the percent of the population above 65 years as our age-related parameter. The variation of Covid-19 cases with the stated parameter is shown in figure 2.

**Figure 2.**
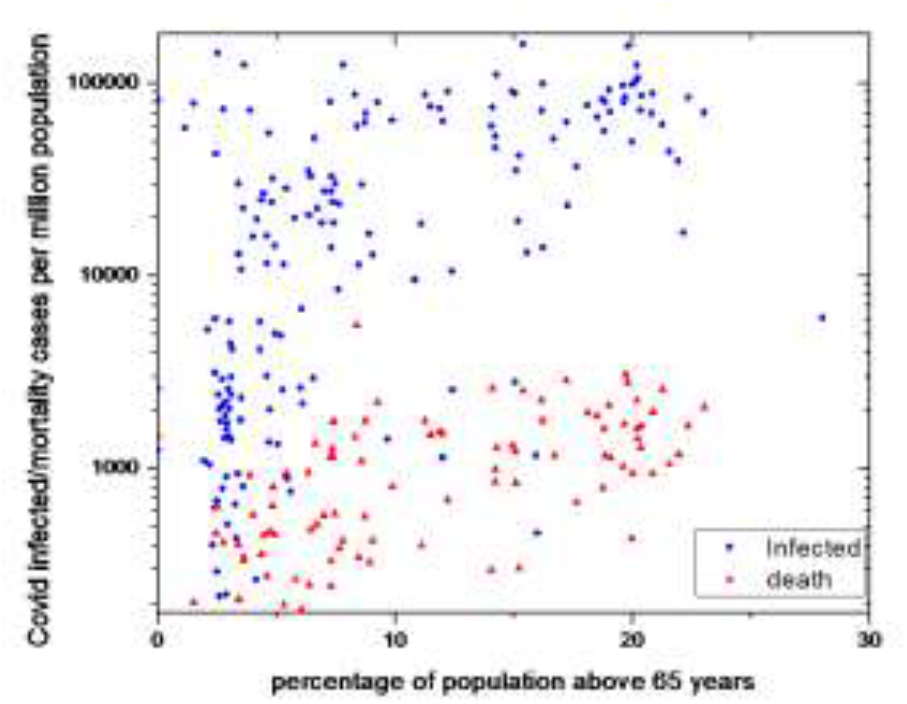
Variation of the Covid-19 infected/mortality cases per million population with the percentage of population above 65 years.

From a correlation analysis we found that the unweighted-correlation coefficient between Covid-19 infected and mortality cases per million population with the percent of the population above 65 years of the countries r = 0.56 and 0.55 respectively. The above findings show a strong association of Covid-19 cases with the age structure of a country. However, with the consideration of weightage the correlation coefficient becomes weaker to 0.45 and 0.39 respectively between Covid-19 infected and mortality cases per million population with the percentage of the population above 65 years.

### 3.4 Covid-19 cases and the urban population

Being an infectious disease the Covid-19 is expected to spread rapidly in dense areas. So we explore for any association between Covid-19 cases with the percentage of the urban population of countries. The percentage of the urban population is normally high in economically rich countries. Figure 3 displays the variation of Covid-19 cases with the percentage of the urban population.

**Figure 3.**
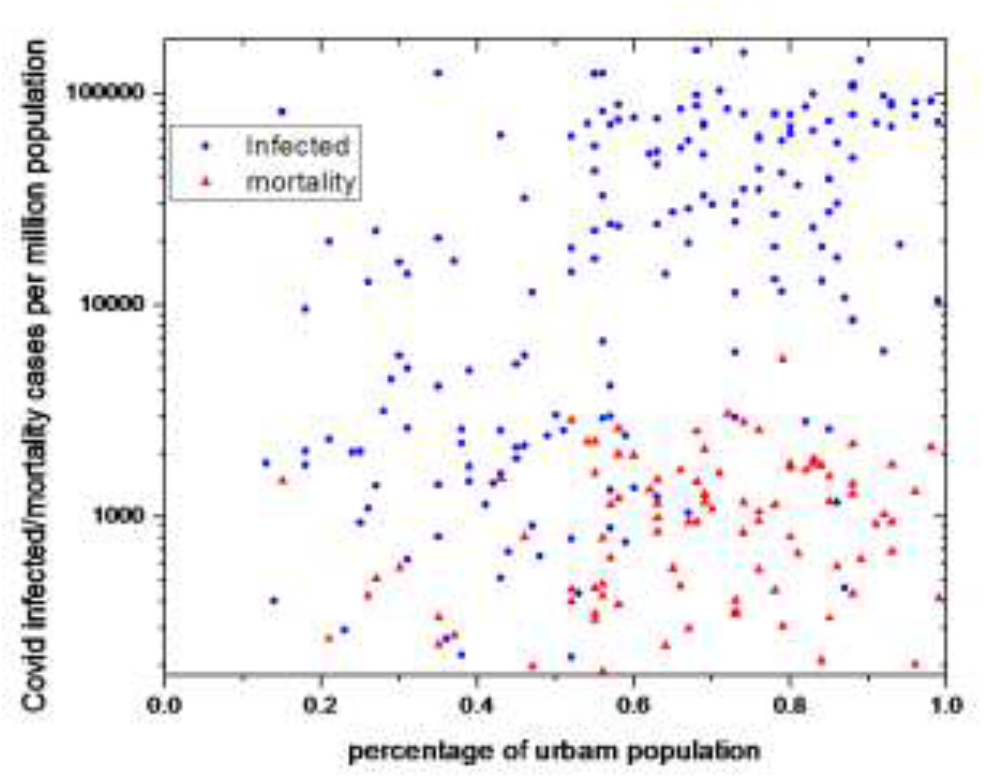
Variation of the Covid-19 infected/mortality cases per million population with the percentage of the urban population of countries.

The correlation coefficients between Covid-19 infected and mortality cases with the percentage of the urban population of countries are 0.48 and 0.38 respectively. However, after consideration of weightage the correlation becomes stronger with r= 0.54 and 0.57 respectively for infected and death cases.

### 3.5 Covid-19 cases and total air passengers

Since Covid-19 is a new disease, initially, it transmits from one continent/country to other mainly through International air travellers. Subsequently, from one city to another within a country mainly through domestic air passengers. We, therefore, explore for any association between Covid-19 cases and total air passengers of countries. To offset the population effect we consider the total air passenger per million population. We have taken the data of total air passengers from the World Bank website. No air passenger data is available for a number of countries like Denmark, Sweden, Central African Republic, etc. Figure 4 portrays the variation of Covid-19 cases in 139 countries with the total air passengers during the year 2019/million population.

**Figure 4:**
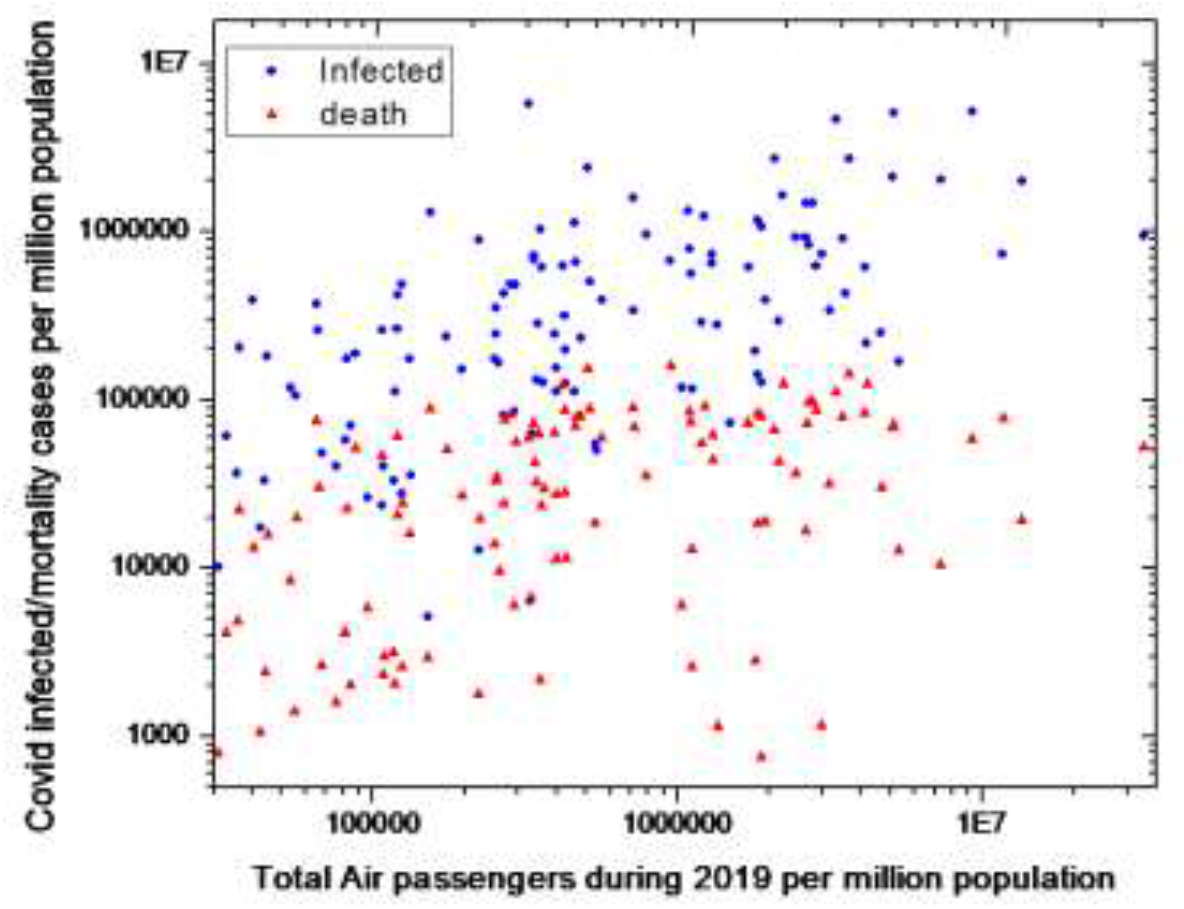
Variation of Covid-19 cases with total air passenger during 2019 per million population of the countries

The correlation analysis gives almost no correlation (r=0.13 and 0.02) between Covid-19 infected and death cases per million people with the total air passenger per million people, Use of the fractional contribution of population of a country to the total population of all (138) countries under consideration as weightage, we, however, found the weighted correlation coefficient r = 0.37 and 0.29 respectively for infected and death cases respectively which shows moderate correlation. The multiple correlation coefficients between Covid-19 cases and the urban population, the percentage of persons of age more than 65 years in the population, along with total air passenger are found 0.61 and 0.57 respectively for Covid-19 infected and death cases.

We have performed linear regression analysis (Eq.4) taking Covid-19 infected (N_i_) and death (N_d_) cases as dependent outcome and the GNI/capita PPP, the percentage of population in urban areas (N_U_), the percentage of persons of age more than 65 years in the population and the total air passenger per thousand person of a country (N_A_) as independent variables and obtained the R^2^_adj_ for the fittings We also fit the data with a power law

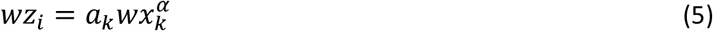

where the subscript i stands for infected and death cases, the subscript k denotes the GNI/capita PPP of a country, the percentage of population in urban areas, the percentage of people above 65 years in population and total air passenger, α is the exponent. Note that we have considered weighted independent and dependent variables taking the fraction of population of a country to the total population of all the countries considered as the weight factor. We find that the linear regression better describes the data for GNI/capita PPP as independent variables whereas the power law better fits the data for other independent variables. The parameters of power law fitting are given in Table 5

**Table 5:** The power law model fitted parameters:

|  | Infected cases |  |  | Mortality cases |  |  |
| --- | --- | --- | --- | --- | --- | --- |
| | A | $\alpha$ | $R^2_{adj}$ | a | $\alpha$ | $R^2_{adj}$ |
| GNI/capita PPP | 8.9±3.9 | 1.2±0.1 | 0.67 | 8.9±3.9 | 1.2±0.1 | 0.67 |
| Percentage of urban population | 0.0068 ± 0.0021 | 0.79±0.04 | 0.77 | 12.9±1.2 | 0.94±0.06 | 0.64 |
| Percentage of people above 65 years | 0.00074 ± 3.9 | 0.85±0.04 | 0.76 | 60.3±4.0 | 0.83±0.06 | 0.59 |
| Total air passenger per 1000 people | 12.4±4.9 | 0.51±0.12 | 0.19 | 3.3±1.2 | 0.44±0.12 | 0.12 |

The coefficient of linear regression for GNI/capita as independent variable are $k=27.65\pm13.02$ and $b=0.46\pm0.02$ with $R^2_{adj}=0.77$ for infected cases whereas for mortality cases $k=0.62\pm0.58$ and $b=0.030\pm0.002$ with $R^2_{adj}=0.67$ .

## Discussion & Conclusion

The present analysis shows that there is a clear positive correlation between Covid-19 infection cases and the economic condition of a country. The mortality cases show only a moderate correlation with the economic condition. Probably because of the better healthcare system in the rich countries the correlation of the Covid-19 death cases with the economic condition is relatively weaker than that of infection cases. Obviously the SARS-CoV-2 virus cannot differentiate economically wellbeing (wealthy) and non-wellbeing people. It has to be any other features or lifestyles of the people of the rich countries that led to large scale spread of Covid-19 infection and consequent mortality there.

The calculated weighted correlation coefficients suggests that there is a positive correlation between Covid-19 cases and median age or the percentage of population above 65 years but the correlation is not very strong. This is consistent with the fact that the total number of elderly people in developing countries are far more than that in rich countries.

The percentage of people living in urban areas is also generally associated with the economic condition of a country. Our analysis shows a strong correlation between Covid-19 cases and the percentage of people living in the urban areas of countries. This implies that the people of rural areas are significantly less affected by the Covid-19 which is not unexpected owing to their somewhat isolation from the rest. Since the above analysis is indicating that the rural areas are largely unaffected by Covid-19 till now, there is also a possibility of some under-reporting from rural areas because of mainly poor relevant infrastructure. However, the rate of positivity analysis in section 3.1 indicates that the percentage of under-reporting is unlikely to be very high.

The large-scale urbanization of rich countries alone cannot be the main cause of high Covid-19 cases in rich countries. Nearly 55% of the world population today live in urban areas. The percentage of people in urban areas (less isolated), a high percentage of people above 65 years and the frequent movement of people leading to diffusion of coronavirus together appear to be the dominant cause for high Covid-19 cases in rich countries. Note that the total air passenger just gives an indication of mixing up of people of different regions but it certainly does not reflect the local mixing up of the people.

The uneven distribution of Covid-19 cases among countries can well be explained in terms of isolation and mix-up of people. The residents of a European or a South American country in general move frequently to other European/South American countries. This leads to continuous diffusion of Covid-19 infection from a highly infected zone to a less infected zone. Because of their large population, such diffusion of Covid-19 infection also happens in India and the USA. India is comparatively less infected than the USA or the European countries because the dominating portion of its population lives in rural areas and the mixing of people of different regions and other countries is also comparatively low. Bangladesh and Pakistan have lower cases of Covid-19 than India because of their relatively smaller population resulting in lower mixing. For instance, West Bengal is a state of India with having a population of around 100 million which is geographically located adjacent to Bangladesh. The Covid-19 cases are higher in West Bengal (infected 1432019 and deceased 16362 as of 7^th^ June 2021) compared to Bangladesh (infected about 0.81 million, deceased 12839) whereas the population of Bangladesh is about 1.6 times higher. Being a part of India, a good fraction of the people of West Bengal travel/visit/migrate different parts of India and vice versa. Such a feature is absent for Bangladesh. Though the novel coronavirus first burst out in China it appears that they could successfully able to isolate the Covid-19 infected regions from the rest of the country. The Covid-19 cases are significantly lower in Australia, New Zealand which are quite isolated and the countries take measures to limited diffusion by restricting International travel, etc. The rate of Covid-19 cases is comparatively low in the countries from Africa, South Asia, and Western pacific from the rest but the rate of Covid-19 cases is one of the highest in the world in the Maldives, a South Asian country. The population of Maldives is 549365 whereas the total tourist arrival in the Maldives in 2020 was 555399 (https://edition.mv › Maldives). Seychelles (Africa) is another similar example where the number of visitors is much higher than its population. The success of Lockdown in containing the Covid-19 cases also support the isolation-diffusion mechanism.

## Conclusion

The world Covid-19 data till 18^th^ June 2021 confirms that the Covid-19 cases are still substantially higher in countries with strong economic conditions compare to the countries of weaker economic position. The present analysis suggests that the lower urban population, limited movement of people, along with lower median age appear the main causes for low Covid-19 cases in those regions. The contribution of under-reporting for lower Covid-19 infected cases in a majority of African, South Asian, and Western-Pacific countries seems not substantial.

The lower-income countries are usually found to be affected much more compared to high-income countries by most of the other infectious diseases like malaria, tuberculosis, typhoid, diphtheria, etc. So Covid-19 is unlikely to be an exception. Since Covid-19 is a new disease, initially it spreads from one continent/country to another mainly through International air traveller who is usually economically strong. Subsequently, the disease spreads to their close community and then to larger people through the cascading process. It seems that the disease is slowly penetrating the rural areas and economically weaker section starts to get infected. It may reach a homogeneous state in the future which will be dangerous for a larger section of the population of the world both economically and physically. If the Covid-19 pandemic eventually becomes an endemic disease nothing like that. Alternatively, It will be great if the vaccination works well though till now the situation is not very straightforward owing to the fast mutation of coronavirus. Hence it is important to frame optimum strategies for isolation and low diffusion to survive against Covid-19 maintaining the prevailing economic activities as much as possible in case the other strategies do not work.

## Data Availability

We have taken the world Covid-19 cumulative infection and death cases data from the WHO website,. Other data are taken from World Bank website and from Worldometer.

